# Environmental surveillance for *Salmonella* Typhi as a tool to estimate the incidence of typhoid fever in low-income populations

**DOI:** 10.1101/2021.05.21.21257547

**Authors:** Christopher B. Uzzell, Catherine M. Troman, Jonathan Rigby, Venkata Raghava Mohan, Jacob John, Dilip Abraham, Rajan Srinivasan, Satheesh Nair, John Scott Meschke, Nicola Elviss, Gagandeep Kang, Nicholas Feasey, Nicholas C. Grassly

## Abstract

**Background:** The World Health Organisation recommends prioritised use of recently prequalified typhoid conjugate vaccines in countries with the highest incidence of typhoid fever. However, representative typhoid surveillance data are lacking in many low-income countries because of the costs and challenges of diagnostic clinical microbiology. Environmental surveillance (ES) of *Salmonella* Typhi in sewage and wastewater using molecular methods may offer a low-cost alternative, but its performance in comparison with clinical surveillance has not been assessed.

**Methodology/Principal Findings:** We developed a harmonised protocol for typhoid ES and its implementation in communities in India and Malawi where it will be compared with findings from hospital-based surveillance for typhoid fever. The protocol includes methods for ES site selection based on geospatial analysis, grab and trap sample collection at sewage and wastewater sites, and laboratory methods for sample processing, concentration and quantitative PCR to detect *Salmonella* Typhi. The optimal locations for ES sites based on digital elevation models and mapping of sewage and river networks are described for each community and their suitability confirmed through field investigation. We will compare the prevalence and abundance of *Salmonella* Typhi in ES samples collected each month over a 12-month period to the incidence of blood culture confirmed typhoid estimated from cases recorded at referral hospitals serving the study areas and community surveys of healthcare seeking for individuals with fever.

**Significance:** If environmental detection of *Salmonella* Typhi correlates with the incidence of typhoid fever estimated through clinical surveillance, typhoid ES may be a powerful and low-cost tool to estimate the local burden of typhoid fever and support the introduction of typhoid conjugate vaccines. Typhoid ES could also allow the impact of vaccination to be assessed and rapidly identify circulation of drug resistant strains.

**Author Summary:** Typhoid fever is a bacterial enteric infection that remains a significant health burden to the 6.4 billion people living in low- and middle-income countries. The WHO recently prequalified highly immunogenic typhoid conjugate vaccines (TCVs) and recommended that the introduction of public vaccination programmes should be based on an understanding of the local epidemiology of infection. However, representative data on the community level describing incidence of microbiologically confirmed typhoid cases are limited, reflecting the costs and challenges of implementing blood culture for *Salmonella* Typhi (*S*. Typhi). Therefore, cost-effective alternative means of community surveillance are required to assess overall burden of disease.

We developed a protocol for typhoid environmental surveillance, including methods for the identification of suitable sampling sites, sample collection and laboratory processing. The detection of *S*. Typhi in wastewater and sewage samples will be examined against the outcomes of hospital-based surveillance for typhoid fever with healthcare utilisation surveys to evaluate and validate the use of ES as an effective means of assessing community level incidence of typhoid. The findings of this study will inform potential wider implementation of ES for typhoid to support the introduction of TCVs and monitor their impact.

## Introduction

Typhoid fever is caused by the human-restricted pathogen *Salmonella enterica* subspecies *enterica* serovar Typhi (*S*. Typhi). Recent estimates suggest that approximately 11 million new typhoid infections occur annually, resulting in approximately 117,000 deaths [1]. Incidence varies geographically with >90% of cases in south and southeast Asia and sub-Saharan Africa, where access to water, sanitation and hygiene (WASH) infrastructure is limited [2,3]. Typhoid is especially prevalent in urban, low socio-economic settings where overcrowding and unhygienic living conditions are common [4,5]. Incidence is typically lower in rural areas, although recent findings in rural or peri-urban sites describe a higher burden than previously appreciated [6,7]. Whilst increasing access to safe drinking water, improved sanitation and antibiotics have helped reduce the incidence of typhoid fever, its persistence in low-income countries and the emergence of antibiotic resistance, including extensively resistant (XDR) strains, are a major concern [8,9].

The isolation of *S*. Typhi by culture remains the gold standard method of detecting typhoid both in clinical practice and for the purposes of typhoid surveillance [10]. However, microbiological culture is time-consuming and resource intensive, and therefore has been difficult to sustain in low-and-middle-income countries (LMICs) where typhoid fever is endemic [11]. Consequently, the Widal test, based on seroconversion to surface (O) and flagellin (H) antigens, is still commonly used to diagnose typhoid, despite its low sensitivity and specificity [12,13]. Currently there are no point-of-care diagnostics that are sufficiently accurate to replace blood culture [14].

By the end of 2020, two Typhoid Conjugate Vaccines (TCV) had been prequalified by the World Health Organisation (WHO) and recommended for use to control typhoid fever with their introduction prioritised in countries with the highest incidence of typhoid disease or a high prevalence of antimicrobial resistant *S*. Typhi [15]. However, limited surveillance capacity means that there is a shortage of reliable country-specific data on the burden of typhoid fever in LMICs. Therefore, alternative strategies are required to better understand the incidence and epidemiology of typhoid fever in resource poor areas, to inform future TCV roll-out and its likely cost-effectiveness [16]. One alternative to blood culture based febrile illness surveillance is to undertake environmental surveillance (ES) whereby pathogens circulating in a population are monitored by testing environmental samples (sewage and wastewater) for their presence. ES is dependent on detecting disease-causing microbiological agents shed into the environment through urine or faeces regardless of the clinical manifestations experienced by the patient. The primary advantage of ES is its relatively low cost and potential for high sensitivity, given the aggregation of faecal material from a large population sample. As a result, it is capable of detecting community-wide subclinical (*i*.*e. “silent”*) disease transmission [17] which contributes to maintenance of *S*. Typhi in the human population.

In recent years, ES has been used to support the Global Poliovirus Eradication Initiative [18,19]. This has stimulated interest in the expanded application of ES for diverse pathogens including *S*. Typhi and more recently SARS-CoV-2 as a complementary approach to clinical and diagnostic surveillance [20,21]. Detection of *S*. Typhi in environmental samples has recently been used to investigate community transmission dynamics and risk factors for typhoid fever in Bangladesh [22], Nepal [23], Democratic Republic of Congo [24] and Nigeria [25]. Retrospective analysis of environmental samples following identified outbreaks of typhoid may provide valuable information regarding the scale and likely source of the infection [26]. Moreover, it has been postulated that prospective ES could be used to evaluate the impact of TCV introduction in the absence or in addition to blood culture surveillance and as a cost-effective approach to guide catch-up TCV vaccination programs [27]. However, the relationship between environmental detection and disease incidence remains unclear as few studies have sought to conduct typhoid ES in communities with ongoing surveillance for typhoid cases.

Here we describe a systematic approach to the design and implementation of typhoid ES, which will begin in India and Malawi in 2021. The performance of ES will be evaluated through comparison of the prevalence and quantity of *S*. Typhi detected in ES samples with typhoid fever incidence inferred from the number of culture-confirmed typhoid cases at local hospitals and health care utilisation surveys. This will indicate the utility of typhoid ES as an affordable tool to determine the incidence of typhoid fever in the community, and the need for vaccination and WASH interventions. We describe the study design for assessing the performance of typhoid ES; develop a protocol for the identification of suitable ES site locations using a remote, geographic information systems (GIS) based framework and local demographic data; outline field-based sample collection, handling, and processing protocols; and describe the laboratory methods for direct detection of *S*. Typhi in ES grab and trap samples.

## Methods

### Study design

The locations for this study will be the central area of Vellore, India and the city of Blantyre, Malawi. Study boundaries are defined as those areas covered by hospital-based blood culture surveillance and planned healthcare utilisation surveys. In Vellore, this corresponds to the study area originally identified by the Surveillance of Enteric Fever in India (SEFI) program [28] and represents an area of 16 km^2^. In Blantyre, this includes the entire municipality, consisting of the central urban districts and surrounding areas, representing 214 km^2^. We will compare the prevalence and abundance of *S*. Typhi in ES samples between these study locations and within locations over time and space during a 12-month period. We will also assess the association between the prevalence and abundance of *S*. Typhi in ES samples and the incidence of culture-confirmed typhoid fever cases in ES site catchment populations as reported at local hospitals.

The estimated incidence of typhoid fever among children in Vellore during 2017-2020 was about twice that for Blantyre during a similar time period 2016-2018 (approximately 2% vs 1% per year of observation for children <15 years old) [29]. Pilot ES surveillance data from Vellore based on direct molecular detection methods using either grab samples (bag-mediated filtration) or trap samples (Moore swabs), detected *S*. Typhi in 14% of all samples. Similar data are not yet available from Blantyre. If typhoid ES is to give a reasonable indication of the incidence of typhoid fever, we might expect the prevalence of *S*. Typhi in ES samples in Blantyre to be about half that observed in Vellore (*i*.*e*. 5-7% vs 14%). To detect this difference in proportions with 80% power we would need to sample between 26 and 47 ES sites with monthly sample collection in each location, using a mixed-effects logistic regression model to allow for clustering of detection by site (the intraclass correlation coefficient in Vellore was 0.08 for the pilot data) [30]. We therefore decided to select 40 ES sites at each location and to sample every month. We will also compare the detection (yes/no) and abundance (genome copies) of *S*. Typhi at each site over time with the incidence of typhoid fever in the associated catchment populations using mixed-effects logistic and linear regression models, respectively. We did not conduct formal power calculations for these analyses because of uncertainty in underlying effect sizes and variance in study measurements.

### ES site selection

The two study locations used for this project are not served by formal, centralised wastewater networks. Instead, waste material and sewage are deposited and transported via either an artificial open above-ground shallow drainage network (Vellore), or an urban freshwater river system that is contaminated directly or through emptying of the contents of pit latrines (Blantyre). Digital geospatially referenced datasets representing the spatial distribution of the wastewater networks were used for the ES site selection process. Within a GIS-based framework, all wastewater confluence points (*i*.*e*. locations where 2 or more channels converge) present throughout the full extent of the 2 study areas were automatically identified and geographic coordinates were calculated and stored. Results were manually checked, and all duplicate or erroneous records were removed. The remaining locations represented the initial list of potential candidate ES sites for each study site.

Freely available elevation data (Advanced Land Observing Satellite Digital Elevation Model (DEM) 12.5 m resolution) was obtained for each study area from the Alaska Satellite Facility (ASF) Distributed Active Archive Center (DAAC). Using the standardised AGREE watershed delineation approach [31,32], whereby the DEMs are hydrologically reconditioned using the geospatially referenced network shapefiles (*i*.*e*. flow enforcement via stream burning), new topographic datasets were created and used to generate geographical catchment areas for each candidate ES site location. Candidate ES locations that generated catchments which did not capture areas contained within the study area boundaries were removed. Of those retained, a duplicate set of ‘digitally clipped’ catchments were produced whereby the full extents were geospatially intersected with the specific study area boundaries. Depending on data availability, the WorldPop [33] or High-Resolution Settlement Layer (HRSL) [34] datasets were used to estimate catchment populations and calculate population density estimates for both the full catchments and those spatially clipped to the study areas. Digital land use and human population datasets indicated key differences in the environmental composition between the two study areas. Specifically, Vellore consists exclusively of medium to high population density urban environments, whilst the larger Blantyre study site exhibits a variety of land use types, including agricultural lands, natural grassland and forested areas in addition to both peri-urban and urban areas characterised by a wide range of residential settings (*e*.*g*. informal, traditional and permanent dwellings). To harmonise the study design and improve consistency and comparability between the 2 locations, we only retained those candidate ES sites throughout the Blantyre study area that produced catchments consisting mostly of medium and high population density urban areas. All other ES sites which did not meet this requirement were deleted.

Each study area was divided into an array of equal size rectangular cells (*i*.*e*. fishnet) used to generate a sampling grid. Due to the differences in areal extents between the two study areas, different size grid networks were selected to ensure appropriate resolution for each location: 500 m and 75 m for Blantyre and Vellore, respectively. The number of candidate ES sites per grid cell was calculated and for each sample cell that contained multiple potential ES locations the downstream site was retained whilst all others were deleted. However, if two or more ES sites in the same grid cell were situated on different river streams or drainage network channels, therefore generating independent catchments, then both/all were retained.

Finally, to ensure that the finalised list of ES sites for each location represented a wide range of catchment sizes, candidate ES sites were stratified by catchment population estimates and categorised into 3 approximately equal size groups (terciles) representing small, medium and large catchment populations. In order to maximise detection sensitivity, ES sites with the greatest population density estimates for each catchment size category were identified and retained as priority ES sites. Once finalised, the full lists of priority candidate ES sites were visited by field-based teams to undertake detailed site descriptions and assess their overall suitability for inclusion in the study based on accessibility, perceived quality of sampling location (*e*.*g*. adequate depth and flow of wastewater) and personnel safety concerns. Sites that were deemed unsuitable where removed. Of the suitable locations, a total of 40 sites for each study area were confirmed as the project ES sites. Where available, previously obtained data during initial pilot studies were used to help guide the decision-making process. During this iterative review, it was considered important to retain broad spatial coverage throughout both study areas and maintain a wide distribution of catchment sizes as determined by population estimates.

### ES sample collection

Following the recommendations of the BMGF Typhoid Environmental Surveillance Expert Advisory Committee (EAC), wastewater/sewage samples will be collected using two separate collection procedures: trap sampling using Moore swabs (*i*.*e*. gauze pad) and grab samples with membrane filtration. Moore swabs made of hospital gauze will be prepared following the protocol described by Sikorski and Levine [35]. At field ES sites, swabs will be fixed in place using wire or twine tied to a fixed object or stone and suspended in flowing wastewater or sewage for 48-72 hours. Upon retrieval, Moore swabs, including their attachment line, will be placed in containers containing 450 ml of Universal Pre-Enrichment (UPE) broth (BD Difco, Fisher Scientific) [36]. For the grab samples, wastewater/sewage samples will be collected using standard sample collection equipment (*e*.*g*. bucket and rope, long handled dipper or similar) used to fill a 1 litre container. Once collected, all sample containers (Moore swab and grab samples) will be disinfected externally and placed in an ice box for immediate return to the laboratory within 4 hours of retrieval. Sample containers will be labelled with sample ID, date, sample type (*e*.*g*. sewage or wastewater) and initials of personnel collecting the sample. All other sampling equipment will be wiped down with disinfectant. Suitable personal protection equipment (PPE) (including gloves, lab coat, glasses, and face mask) will be donned upon arrival at the sampling site and GPS coordinates will be recorded using handheld GPS device. Each quarter, wastewater physicochemical properties will also be measured at the site (including temperature, pH, oxidative reduction potential (ORP), dissolved oxygen, total suspended solids (TDS), salinity and turbidity) using a water quality probe following a published protocol [17]. At the first visit field staff will also collect information on local infrastructure and ES site characteristics using an electronic questionnaire.

### Blood culture surveillance

Information on the number, age, and place of residence for blood culture confirmed typhoid cases recorded at hospitals serving the study communities during the study period will be extracted from the electronic records. In Vellore, the Christian Medical College (CMC) hospital, a 1,721 bed tertiary care centre, and its two satellite facilities, the Low Cost Effective Care Unit and the CHAD hospital, serve the study area along with the government-run Pentland Hospital. We will use health seeking survey instruments to determine the probability that a typhoid case seeks care at one of these participating hospitals. Age-appropriate blood volumes (3 ml in <1 year; 5 ml in 1 to 15 years; 8 to 10 ml >15 years) will be obtained from those hospitalised with an acute febrile illness and culture for *S*. Typhi done using the automated BacT/ALERT system (bioMérieux, France) at the CMC Vellore microbiology department.

In Blantyre, the Queen Elizabeth Central Hospital (QECH) serves the study area and surrounding districts. Blood will be sampled from paediatric patients presenting with non-specific febrile illness, who test negative for malaria, are severely ill with suspected sepsis, or fail initial malaria treatment and remain febrile, and from all febrile adult patients admitted. 2-4 mL of blood will be taken for culture from children (aged younger than 16 years) and 7-10 mls from adults under aseptic conditions. All blood samples will be cultured using the automated BacT/ALERT system (bioMérieux, France).

Bottles that flag as positive after incubation will have Gram stain done and Gram-negative bacilli will be identified using API biochemical testing (bioMérieux, France). Salmonellae, including *S*. Typhi will be typed using agglutination with serotype-specific antisera according to the White-Kauffman-Le Minor scheme [37].

### ES laboratory methods

On arrival at the laboratory, Moore swabs will be incubated at 37°C for 24 hours in the containers of UPE to enrich bacteria trapped in the swab. 40 ml will be filtered in two aliquots of 20 ml under a vacuum and the filter discs cut into strips and stored at −20°C until DNA extraction. Grab samples for membrane filtration will be pre-filtered using a coffee filter, then filtered under a vacuum using up to five filter discs (0.45 µm) which will then be eluted using 10 ml Ringers lactate solution. 1 ml of the Ringers lactate solution will then be centrifuged, and the resulting pellet stored at −20°C until DNA extraction. These methods have shown to work adequately for processing Moore swabs and grab samples and for downstream detection of *S*. Typhi [36].

DNA extraction will be carried out using the QIAamp PowerFecal Pro kit (Qiagen) following the manufacturer’s protocol, eluted into 50 µl, and stored at −20°C. A commercially available spike-in control will be used to assess success of DNA extraction and PCR inhibition (qPCR DNA Extraction and Inhibition Control, Eurogentec). The quantitative (q)PCR targets three genes in the *S*. Typhi genome: *staG, ttr*, and *tviB*. Of these, *staG* and *tviB* in combination have been shown to detect *S*. Typhi [38,39], whilst *ttr* should give a positive result for all *Salmonella* species [38]. Primers and probes targeting HF183, a frequently used marker gene derived from Bacteroides 16S rRNA, have been selected for use as a positive control to detect human faecal contamination of the samples [40]. Primer and probe sequences to be used in this study are detailed in Table 1.

**Table 1.**
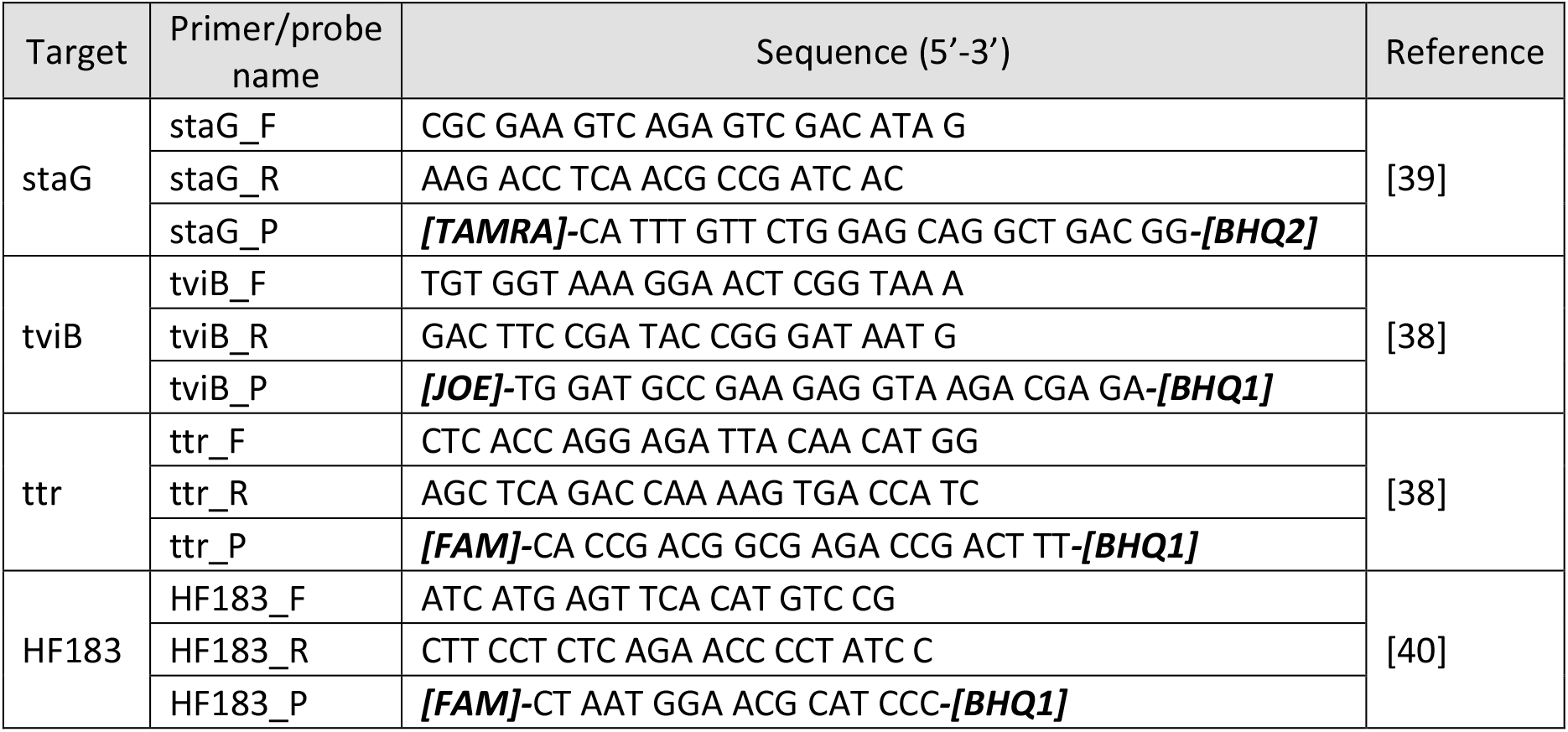
Sequences for the primers and probes to be used in this study. Fluorescent dyes and quenchers on the probes are in bold.

Reactions can be carried out in a multiplex or as single reactions and will be carried out in a final volume of 25 µl using a qPCR mastermix with a ROX reference dye for normalisation (multiple mixes will be evaluated). Primers and probes will be included at a final concentration of 0.5µM and 0.1µM respectively for *tviB, staG* and HF183, and *ttr* primers and probe at 0.25µM and 0.5µM. Cycling conditions for all qPCR reactions will be as follows: 50°C for 2 minutes then 95°C for 15 minutes followed by 40 cycles of 95°C for 30s, 60°C for 30s and 72°C for 30s. The Ct values for each gene target will be assessed to decide whether a sample is positive for *S*. Typhi. A standard curve for each target will be produced by running a qPCR on a dilution series of gBlocks Gene Fragment DNA standards (IDT) in water.

### Statistical analysis

We will compare the prevalence of *S*. Typhi in ES samples between Vellore and Blantyre using a mixed-effects logistic regression model with a random effect on the intercept by ES site to allow for repeated measures. In each location, mixed-effects logistic regression and binomial regression will be used to investigate the association of *S*. Typhi detection with ES site characteristics including the physicochemical properties of the wastewater/sewage, local infrastructure, climate variables, catchment population size and characteristics, including the number of blood culture confirmed typhoid cases among residents within the catchment area. We will also conduct exploratory analyses of the spatial distribution of *S*. Typhi detection, looking for hotspots for transmission and their association with local population characteristics.

## Results

### Site Selection

The geographic distribution of both candidate and field-based confirmed ES sites for the Blantyre and Vellore study areas is shown in Fig 1A and Fig 1B, respectively, and summary statistics for both the full and spatially intersected catchments are presented in Table 2. Full details can be found in the supplementary material (S1 Table) For Blantyre, an initial list of 93 unique priority candidate locations were identified using the GIS-based site selection approach. Candidate ES sites were relatively well distributed throughout the study area; however, the majority were located throughout the northern and eastern quarters of Blantyre, consistent with the distribution of densely populated areas served by the Mudi and Lukhubula rivers which drain to the southwest and northwest, respectively. Hydrological modelling indicates that catchments for candidate ES site ranged from approx. 0.1 km^2^ to 21.2 km^2^ (mean: 5.1 km^2^). Moreover, catchment population estimates varied between approx. 400 to over 107,000, stratified into 3 equal number groups (terciles; n31): small <9,000; medium 9,000 – 35,000; and large >35,000. Mean population density for all 93 catchments was 8,161 inhabitants/km^2^ (range: 3,550 – 30,268 inhabitants/km^2^), remaining relatively consistent between the population stratified groups with estimates of 9,086 inhabitants/km^2^, 8,056 inhabitants/km^2^ and 7,340 inhabitants/km^2^ for the small, medium and large population terciles, respectively. Most of the ES sites catchments overlapped with the study area, such that the mean extent and population of the full catchments and those clipped to the study area were similar.

**Fig 1.**
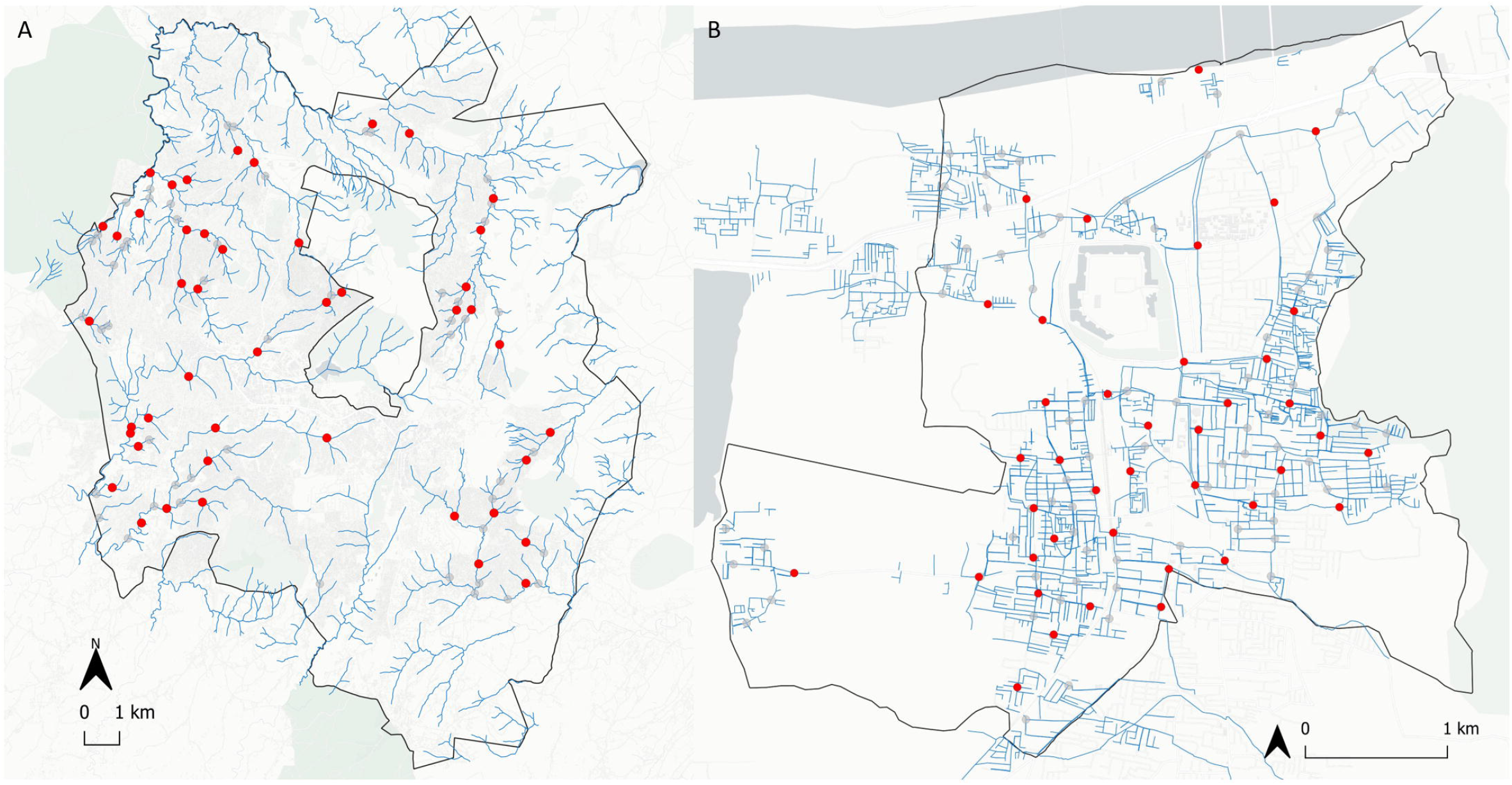
Geographic distribution of ES sites for Blantyre, Malawi (A) and Vellore, India (B). Candidate and confirmed ES sites are shown as grey and red dots, respectively. Blue lines represent the spatial distribution of wastewater networks. Maps were plotted using QGIS 3.10 using base map tiles sourced from CartoDB (using data by OpenStreetMap made available under the Creative Commons Attribution (CC BY) 4.0 license).

**Table 2.**
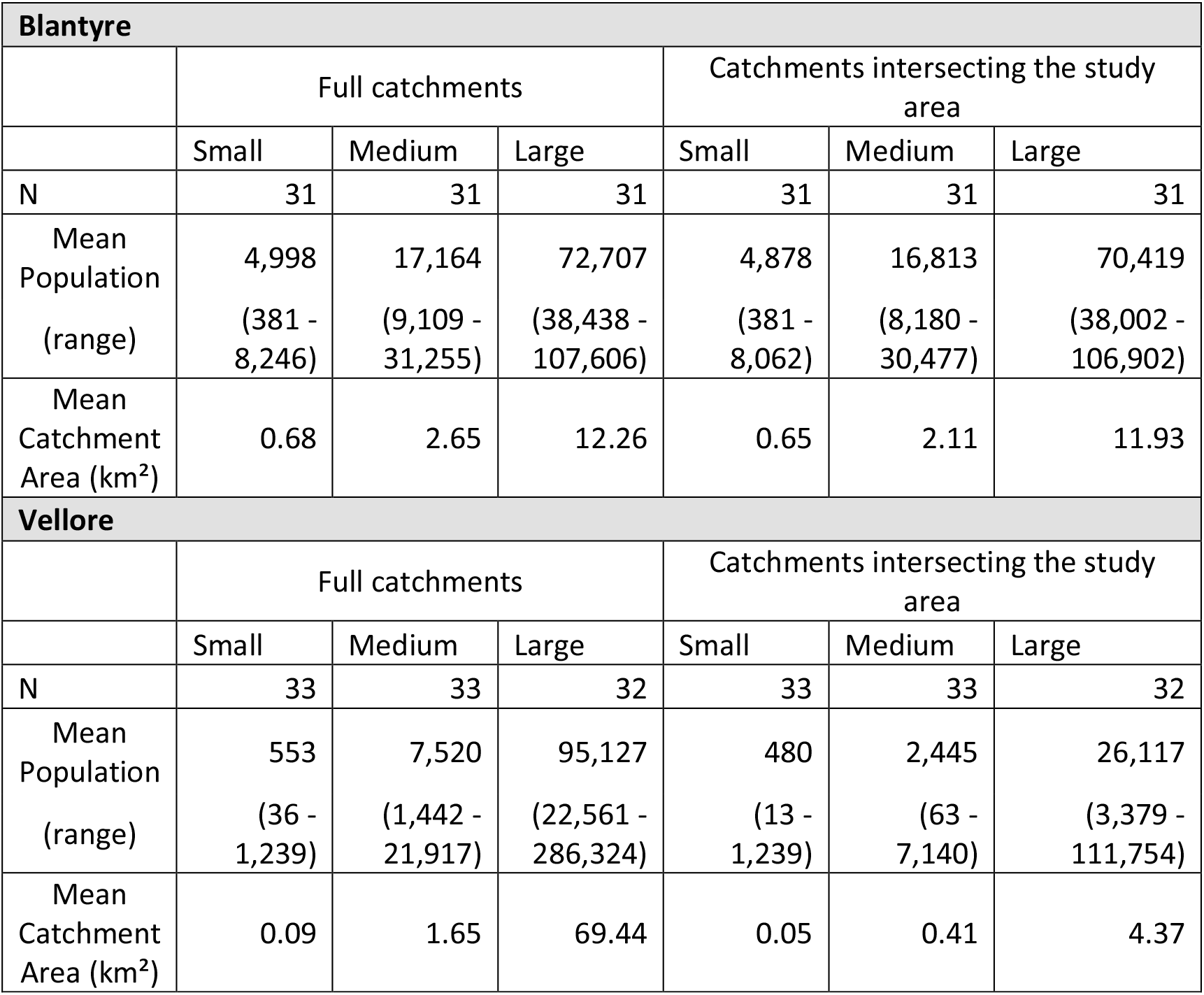
Summary area and population statistics for ES catchments by tercile in Blantyre and Vellore for the full catchments and those overlapping the study area.

| **Blantyre** |  |  |  |  |  |  |
| --- | --- | --- | --- | --- | --- | --- |
|  | Full catchments |  |  | Catchments intersecting the study area |  |  |
|  | Small | Medium | Large | Small | Medium | Large |
| N | 31 | 31 | 31 | 31 | 31 | 31 |
| Mean Population<br>(range) | 4,998<br>(381 - 8,246) | 17,164<br>(9,109 - 31,255) | 72,707<br>(38,438 - 107,606) | 4,878<br>(381 - 8,062) | 16,813<br>(8,180 - 30,477) | 70,419<br>(38,002 - 106,902) |
| Mean Catchment Area (km<sup>2</sup>) | 0.68 | 2.65 | 12.26 | 0.65 | 2.11 | 11.93 |
| **Vellore** |  |  |  |  |  |  |
|  | Full catchments |  |  | Catchments intersecting the study area |  |  |
|  | Small | Medium | Large | Small | Medium | Large |
| N | 33 | 33 | 32 | 33 | 33 | 32 |
| Mean Population<br>(range) | 553<br>(36 - 1,239) | 7,520<br>(1,442 - 21,917) | 95,127<br>(22,561 - 286,324) | 480<br>(13 - 1,239) | 2,445<br>(63 - 7,140) | 26,117<br>(3,379 - 111,754) |
| Mean Catchment Area (km<sup>2</sup>) | 0.09 | 1.65 | 69.44 | 0.05 | 0.41 | 4.37 |

For Vellore, a total of 98 candidate ES site locations were initially identified. The bulk of the sites were located within the study boundary and spatially consistent with those areas better served by the artificial open drainage sewer network. Overall, the mean area for the full unprocessed catchments was 22.5 km^2^, however, this masks significant variability and a highly skewed distribution of catchment sizes ranging from <1 km^2^ to 175 km^2^ (median 0.81 km^2^) with many catchments extending well beyond the study boundary. Similarly, catchment population estimates varied considerably from <50 to over 280,000. Using the tercile stratification, small medium and large class catchments were based on approximate population thresholds of <1,300, 1,300 – 22,000 and >22,000, respectively. Catchment extent and population metrics were significantly altered when using the processed catchments that spatially intersected with the study boundary. Using these data, the maximum catchment size was 15.6 km^2^ (approx. 94% of the study area), representing an estimated population of approximately 111,750. Moreover, mean population density estimates using the cropped catchment extents remained relatively consistent, ranging from 6,863 inhabitants/km^2^ to 5,534 inhabitants/km^2^ for small and large category catchments, respectively. These refinements were not required for Blantyre as the entire urban area was selected as described above.

Candidate ES site locations underwent a significant review process and following field-based site investigations and detailed discussions between study partners in each study location, the proposed lists of candidate sites were used to identity and confirm the 40 ES sites to be retained for the study. ES sites confirmed for inclusion in the study are presented in Fig 1a and Fig 1b.

## Discussion

The support for widespread TCV introduction by the WHO Strategic Advisory Group of Experts (SAGE) on immunisation [41] and approval of funding by Gavi offer hope in the fight to reduce morbidity and mortality from typhoid fever. Introduction of TCV into a country’s vaccine schedule should be informed by country-specific data on the baseline burden of typhoid. However, these data are typically lacking because of limited implementation of hospital-based blood culture surveillance or of community-based surveillance for acute febrile illness. Moreover, these surveillance methods may lack sensitivity in communities with a high uptake of effective antibiotics or where formal healthcare is not always sought. ES may therefore offer a low-cost, sensitive alternative or supplement to clinical surveillance, providing critical information on the burden of typhoid that will support TCV introduction [42]. However, few studies have investigated typhoid ES and therefore, uncertainties regarding how best to effectively design and implement such a surveillance strategy remain.

To address these gaps, we describe the design of a study to detect *S*. Typhi in environmental samples and correlate these findings with contemporaneous hospital-based blood culture surveillance serving the same communities over a 12-month period. This will provide timely information to help ascertain the technical feasibility and reproducibility of typhoid ES and its utility in determining the local incidence of typhoid fever. It will also inform our understanding of the spatiotemporal patterns of *S*. Typhi transmission within the community and the optimal ES site characteristics for *S*. Typhi detection.

The relatively low cost and scalability of ES means it can also be expanded to cover large geographic areas that include a range of settings (*i*.*e*. rural, urban and peri-urban) therefore allowing decision-makers to better understand the epidemiology of infection and develop and deploy an optimum region-specific vaccination programme. Specifically, such information could allow for the more appropriate distribution of TCV relative to the spatial burden of typhoid fever. ES could also be deployed in the event of a suspected outbreak of typhoid fever and used to locate high risk areas for targeted vaccination. Finally, ES may also be valuable in evaluating the impact of TCV following its roll out by monitoring potential reductions in *S*. Typhi prevalence or abundance in environmental samples.

In preparing the current study protocol, several important considerations were made. Firstly, we develop a representative site selection strategy that ensures the findings of the ES can be effectively compared with the incidence of typhoid fever estimated from healthcare utilisation surveys and the number of cases reported by local hospitals serving the community. Secondly, the study design sought to maximise the comparability of results between countries by standardising the methodological approach. Thirdly, we aimed to generate an easily reproducible approach to be adopted as a standardised procedure for future studies. This was particularly important as we plan to expand the proposed study to include a third study site in the Asante Akim North district, Ghana.

We believe that the design of this prospective study has several strengths. First, we anticipate that the inclusion of two distinct, yet comparable study sites characterised by differing environmental conditions will provide an opportunity to elucidate potential differences or similarities in the distribution and abundance of *S*. Typhi and performance of ES between these settings. Second, in ensuring the broad geographic distribution of ES sites at each study area, therefore capturing a wide range of microenvironments, we will be able to investigate how different ES site and catchment characteristics might correlate with the detection of *S*. Typhi. We also acknowledge that the proposed study may be affected by several limitations. First, upon the initial commencement of the study, the program of ES will be geographically restricted to two locations and temporally limited to just one year of active surveillance. Moreover, due to the ongoing global COVID-19 pandemic, we recognise that healthcare utilisation patterns within the study areas are likely to be impacted during the study period. Regarding the Vellore study location, due to the relatively coarse spatial resolution of the DEM used during the hydrological modelling process, coupled with the high level of detail in the wastewater network, there is some uncertainty in the drainage catchments, and therefore population metric estimates, for the proposed ES site locations. However, we anticipate that the collection of ancillary data (such as local hydrologically) during the initial phase of the study, will allow us to refine the catchment modelling process and improve predictive accuracy of catchment wide population estimates. Finally, we recognise that despite significant advancements in molecular detection methods, there remain uncertainties regarding its sensitivity to detect *S*. Typhi in ES samples [26]. For example, molecular detection from complex environmental samples is often hampered by relatively low sensitivity and specificity [35] and therefore may yield false-negative results, thus prompting an underestimation of *S*. Typhi in the environmental samples [43].

Whilst active acute febrile illness surveillance with microbiological confirmation by culture remains the gold standard for estimation of the burden of typhoid fever in a community, such approaches are hugely time and resource demanding. Well designed and sensitive ES of *S*. Typhi may offer a practical and relatively low-cost alternative that could rapidly generate vital information regarding the prevalence of infection. This information could be used by national technical advisory groups and others to determine the likely benefits of TCV introduction and to monitor the impact of vaccination or WASH interventions to tackle typhoid fever.

## Data Availability

All data referenced to in this manuscript are fully available without restriction and available in the Supplementary Information documents.

## Supporting information captions

**S1 Table**. Full area and population statistics for environmental surveillance catchments in Blantyre and Vellore for full catchments and those overlapping the study area.

