## Supplemental Table 1 for "Environmental surveillance for *Salmonella* Typhi as a tool to estimate the incidence of typhoid fever in low-income populations"

**S1 Table.** Full area and population statistics for environmental surveillance catchments in Blantyre and Vellore for full catchments and those overlapping the study area.

|  |  | Full catchments | | | Catchments intersecting the study area | | |
| --- | --- | --- | --- | --- | --- | --- | --- |
| ID | Study Location | Catchment Area (km^2^) | Catchment Population | Catchment Population Density | Catchment Area (km^2^) | Catchment Population | Catchment Population Density |
| B-01 | Blantyre | 0.17 | 1,247 | 7,347 | 0.17 | 1,247 | 7,354 |
| B-02 | Blantyre | 8.86 | 83,490 | 9,424 | 6.96 | 72,366 | 10,403 |
| B-03 | Blantyre | 8.17 | 77,851 | 9,533 | 6.26 | 66,140 | 10,558 |
| B-04 | Blantyre | 0.58 | 3,601 | 6,169 | 0.58 | 3,563 | 6,106 |
| B-05 | Blantyre | 0.30 | 1,592 | 5,317 | 0.30 | 1,592 | 5,319 |
| B-06 | Blantyre | 0.43 | 3,499 | 8,105 | 0.43 | 3,499 | 8,108 |
| B-07 | Blantyre | 6.59 | 72,260 | 10,961 | 4.69 | 62,849 | 13,399 |
| B-08 | Blantyre | 20.24 | 107,606 | 5,317 | 20.11 | 106,902 | 5,315 |
| B-09 | Blantyre | 5.88 | 63,771 | 10,845 | 3.98 | 53,240 | 13,380 |
| B-10 | Blantyre | 16.51 | 65,584 | 3,972 | 15.86 | 65,502 | 4,130 |
| B-11 | Blantyre | 0.55 | 6,866 | 12,375 | 0.55 | 6,866 | 12,379 |
| B-12 | Blantyre | 15.79 | 77,176 | 4,887 | 15.79 | 76,719 | 4,859 |
| B-13 | Blantyre | 3.01 | 19,257 | 6,402 | 2.89 | 19,077 | 6,606 |
| B-14 | Blantyre | 2.86 | 19,293 | 6,747 | 2.74 | 19,076 | 6,964 |
| B-15 | Blantyre | 15.37 | 65,581 | 4,267 | 14.72 | 65,222 | 4,431 |
| B-16 | Blantyre | 4.13 | 21,876 | 5,294 | 1.19 | 20,598 | 17,305 |
| B-17 | Blantyre | 15.18 | 65,399 | 4,308 | 14.53 | 65,060 | 4,477 |
| B-18 | Blantyre | 10.80 | 60,763 | 5,624 | 10.80 | 60,739 | 5,624 |
| B-19 | Blantyre | 2.20 | 19,246 | 8,766 | 2.08 | 18,920 | 9,116 |
| B-20 | Blantyre | 7.37 | 51,007 | 6,918 | 7.37 | 50,760 | 6,886 |
| B-21 | Blantyre | 12.61 | 64,343 | 5,101 | 11.96 | 63,987 | 5,348 |
| B-22 | Blantyre | 2.93 | 16,199 | 5,537 | 0.65 | 15,335 | 23,617 |
| B-23 | Blantyre | 7.23 | 50,529 | 6,988 | 7.23 | 50,225 | 6,948 |
| B-24 | Blantyre | 12.41 | 64,811 | 5,223 | 11.76 | 64,304 | 5,468 |
| B-25 | Blantyre | 1.45 | 10,903 | 7,515 | 1.45 | 10,630 | 7,329 |
| B-26 | Blantyre | 0.09 | 1,925 | 22,243 | 0.09 | 1,906 | 22,023 |
| B-27 | Blantyre | 0.63 | 5,792 | 9,221 | 0.26 | 5,672 | 22,104 |
| B-28 | Blantyre | 1.79 | 17,806 | 9,928 | 1.67 | 17,689 | 10,569 |
| B-29 | Blantyre | 1.94 | 58,829 | 30,268 | 1.71 | 47,722 | 27,852 |
| B-30 | Blantyre | 0.98 | 7,580 | 7,732 | 0.98 | 7,580 | 7,735 |
| B-31 | Blantyre | 1.64 | 17,354 | 10,588 | 1.52 | 17,263 | 11,365 |
| B-32 | Blantyre | 0.75 | 4,784 | 6,353 | 0.75 | 4,560 | 6,058 |
| B-33 | Blantyre | 1.09 | 11,560 | 10,613 | 0.97 | 11,356 | 11,713 |
| B-34 | Blantyre | 0.23 | 2,323 | 9,953 | 0.23 | 2,323 | 9,956 |
| B-35 | Blantyre | 4.42 | 22,092 | 5,002 | 4.42 | 22,092 | 5,002 |
| B-36 | Blantyre | 0.41 | 5,040 | 12,223 | 0.41 | 5,040 | 12,227 |
| B-37 | Blantyre | 1.14 | 16,544 | 14,499 | 0.32 | 14,113 | 43,616 |
| B-38 | Blantyre | 0.85 | 4,353 | 5,148 | 0.31 | 4,328 | 14,036 |
| B-39 | Blantyre | 3.79 | 31,255 | 8,238 | 0.62 | 26,005 | 41,708 |
| B-40 | Blantyre | 0.56 | 10,494 | 18,781 | 0.49 | 10,417 | 21,386 |
| B-41 | Blantyre | 4.30 | 46,658 | 10,841 | 1.11 | 41,139 | 36,995 |
| B-42 | Blantyre | 2.61 | 11,673 | 4,467 | 2.61 | 11,673 | 4,467 |
| B-43 | Blantyre | 0.46 | 8,071 | 17,691 | 0.38 | 7,996 | 20,788 |
| B-44 | Blantyre | 4.57 | 21,696 | 4,749 | 4.57 | 21,204 | 4,643 |
| B-45 | Blantyre | 1.92 | 16,218 | 8,463 | 1.75 | 16,054 | 9,150 |
| B-46 | Blantyre | 2.31 | 11,391 | 4,942 | 2.27 | 11,295 | 4,978 |
| B-47 | Blantyre | 0.18 | 4,472 | 25,397 | 0.16 | 4,021 | 25,405 |
| B-48 | Blantyre | 1.20 | 7,037 | 5,879 | 1.15 | 7,007 | 6,092 |
| B-49 | Blantyre | 0.06 | 381 | 5,966 | 0.06 | 381 | 5,968 |
| B-50 | Blantyre | 0.28 | 1,510 | 5,376 | 0.28 | 1,482 | 5,276 |
| B-51 | Blantyre | 0.93 | 10,763 | 11,596 | 0.89 | 10,719 | 12,005 |
| B-52 | Blantyre | 6.57 | 69,327 | 10,548 | 3.37 | 63,835 | 18,956 |
| B-53 | Blantyre | 1.20 | 8,246 | 6,894 | 1.20 | 8,032 | 6,718 |
| B-54 | Blantyre | 7.54 | 70,159 | 9,304 | 4.33 | 64,425 | 14,864 |
| B-55 | Blantyre | 1.08 | 4,487 | 4,155 | 1.08 | 4,437 | 4,111 |
| B-56 | Blantyre | 1.56 | 7,687 | 4,915 | 1.56 | 7,687 | 4,917 |
| B-57 | Blantyre | 7.34 | 28,573 | 3,891 | 7.32 | 28,570 | 3,904 |
| B-58 | Blantyre | 2.57 | 10,889 | 4,241 | 2.57 | 10,889 | 4,243 |
| B-59 | Blantyre | 2.61 | 10,611 | 4,064 | 2.59 | 10,486 | 4,054 |
| B-60 | Blantyre | 0.26 | 2,380 | 9,335 | 0.26 | 2,380 | 9,335 |
| B-61 | Blantyre | 0.49 | 7,729 | 15,694 | 0.49 | 7,729 | 15,694 |
| B-62 | Blantyre | 1.53 | 10,360 | 6,774 | 1.53 | 10,283 | 6,726 |
| B-63 | Blantyre | 1.40 | 7,672 | 5,464 | 1.40 | 7,672 | 5,466 |
| B-64 | Blantyre | 1.08 | 5,999 | 5,543 | 1.08 | 5,999 | 5,546 |
| B-65 | Blantyre | 2.29 | 18,727 | 8,187 | 2.29 | 18,727 | 8,190 |
| B-66 | Blantyre | 2.93 | 24,775 | 8,465 | 2.93 | 24,685 | 8,437 |
| B-67 | Blantyre | 0.29 | 3,485 | 12,148 | 0.29 | 3,485 | 12,154 |
| B-68 | Blantyre | 12.01 | 57,198 | 4,763 | 11.98 | 56,889 | 4,748 |
| B-69 | Blantyre | 0.42 | 5,428 | 12,781 | 0.42 | 5,342 | 12,583 |
| B-70 | Blantyre | 0.72 | 8,102 | 11,322 | 0.72 | 8,063 | 11,270 |
| B-71 | Blantyre | 1.23 | 16,754 | 13,585 | 1.23 | 16,723 | 13,565 |
| B-72 | Blantyre | 1.51 | 7,570 | 5,021 | 1.51 | 7,374 | 4,893 |
| B-73 | Blantyre | 6.56 | 38,438 | 5,858 | 6.56 | 38,438 | 5,860 |
| B-74 | Blantyre | 19.92 | 98,218 | 4,931 | 19.89 | 98,165 | 4,936 |
| B-75 | Blantyre | 19.44 | 97,702 | 5,026 | 19.41 | 97,491 | 5,023 |
| B-76 | Blantyre | 2.10 | 30,336 | 14,455 | 2.10 | 30,336 | 14,461 |
| B-77 | Blantyre | 4.21 | 14,928 | 3,550 | 4.21 | 14,928 | 3,550 |
| B-78 | Blantyre | 0.92 | 4,540 | 4,939 | 0.92 | 4,468 | 4,880 |
| B-79 | Blantyre | 20.42 | 99,856 | 4,891 | 20.39 | 99,323 | 4,872 |
| B-80 | Blantyre | 7.89 | 54,376 | 6,892 | 7.89 | 54,376 | 6,895 |
| B-81 | Blantyre | 21.22 | 100,697 | 4,746 | 21.22 | 100,697 | 4,746 |
| B-82 | Blantyre | 0.88 | 4,608 | 5,216 | 0.88 | 4,608 | 5,218 |
| B-83 | Blantyre | 1.78 | 9,109 | 5,120 | 1.78 | 9,109 | 5,122 |
| B-84 | Blantyre | 9.18 | 62,371 | 6,797 | 9.18 | 62,371 | 6,797 |
| B-85 | Blantyre | 1.21 | 6,926 | 5,743 | 1.21 | 6,926 | 5,745 |
| B-86 | Blantyre | 9.68 | 66,407 | 6,864 | 9.68 | 66,407 | 6,864 |
| B-87 | Blantyre | 16.56 | 94,035 | 5,679 | 16.56 | 94,035 | 5,679 |
| B-88 | Blantyre | 17.13 | 98,183 | 5,732 | 17.13 | 98,183 | 5,732 |
| B-89 | Blantyre | 1.83 | 14,396 | 7,878 | 1.83 | 14,396 | 7,878 |
| B-90 | Blantyre | 15.03 | 84,957 | 5,652 | 15.03 | 84,957 | 5,654 |
| B-91 | Blantyre | 15.97 | 86,343 | 5,405 | 15.97 | 86,343 | 5,407 |
| B-92 | Blantyre | 2.65 | 13,415 | 5,066 | 0.55 | 13,302 | 24,350 |
| B-93 | Blantyre | 1.91 | 23,580 | 12,346 | 1.30 | 23,556 | 18,108 |
| V-01 | Vellore | 4.17 | 21,863 | 5,248 | 1.00 | 7,140 | 7,106 |
| V-02 | Vellore | 0.05 | 398 | 8,072 | 0.05 | 398 | 8,072 |
| V-03 | Vellore | 0.10 | 543 | 5,692 | 0.10 | 543 | 5,692 |
| V-04 | Vellore | 140.32 | 164,222 | 1,170 | 6.41 | 28,610 | 4,466 |
| V-05 | Vellore | 0.09 | 534 | 6,027 | 0.05 | 406 | 8,889 |
| V-06 | Vellore | 0.04 | 336 | 8,768 | 0.04 | 336 | 8,768 |
| V-07 | Vellore | 0.66 | 4,294 | 6,484 | 0.66 | 4,294 | 6,484 |
| V-08 | Vellore | 0.18 | 1,239 | 6,712 | 0.18 | 1,239 | 6,712 |
| V-09 | Vellore | 26.32 | 53,436 | 2,030 | 3.95 | 31,183 | 7,902 |
| V-10 | Vellore | 26.36 | 54,021 | 2,049 | 3.99 | 31,768 | 7,962 |
| V-11 | Vellore | 0.03 | 153 | 4,415 | 0.03 | 108 | 3,241 |
| V-12 | Vellore | 0.67 | 2,617 | 3,902 | 0.37 | 1,466 | 3,909 |
| V-13 | Vellore | 0.12 | 828 | 6,828 | 0.06 | 398 | 7,000 |
| V-14 | Vellore | 0.50 | 2,950 | 5,863 | 0.50 | 2,950 | 5,863 |
| V-15 | Vellore | 0.03 | 37 | 1,173 | 0.03 | 37 | 1,218 |
| V-16 | Vellore | 3.40 | 18,645 | 5,486 | 0.83 | 6,219 | 7,489 |
| V-17 | Vellore | 0.74 | 3,173 | 4,279 | 0.45 | 2,022 | 4,536 |
| V-18 | Vellore | 2.01 | 9,689 | 4,815 | 0.10 | 643 | 6,591 |
| V-19 | Vellore | 1.89 | 8,712 | 4,618 | 0.02 | 113 | 5,550 |
| V-20 | Vellore | 19.91 | 17,522 | 880 | 0.26 | 826 | 3,172 |
| V-21 | Vellore | 1.42 | 5,824 | 4,096 | 0.07 | 435 | 6,359 |
| V-22 | Vellore | 3.37 | 16,976 | 5,030 | 0.03 | 128 | 4,343 |
| V-23 | Vellore | 0.45 | 671 | 1,503 | 0.34 | 527 | 1,529 |
| V-24 | Vellore | 0.05 | 49 | 967 | 0.02 | 13 | 737 |
| V-25 | Vellore | 0.04 | 36 | 957 | 0.00 | 36 | 7,927 |
| V-26 | Vellore | 0.14 | 196 | 1,443 | 0.09 | 126 | 1,402 |
| V-27 | Vellore | 0.06 | 133 | 2,194 | 0.05 | 107 | 2,099 |
| V-28 | Vellore | 0.20 | 315 | 1,612 | 0.14 | 240 | 1,726 |
| V-29 | Vellore | 0.15 | 949 | 6,393 | 0.02 | 237 | 14,464 |
| V-30 | Vellore | 0.68 | 2,623 | 3,838 | 0.09 | 326 | 3,593 |
| V-31 | Vellore | 0.28 | 1,820 | 6,509 | 0.28 | 1,820 | 6,509 |
| V-32 | Vellore | 4.57 | 25,148 | 5,499 | 1.41 | 10,425 | 7,384 |
| V-33 | Vellore | 5.94 | 36,413 | 6,130 | 2.78 | 21,690 | 7,804 |
| V-34 | Vellore | 2.85 | 13,887 | 4,873 | 0.64 | 3,689 | 5,769 |
| V-35 | Vellore | 0.78 | 3,377 | 4,324 | 0.49 | 2,226 | 4,586 |
| V-36 | Vellore | 3.07 | 15,716 | 5,120 | 0.69 | 4,300 | 6,243 |
| V-37 | Vellore | 4.14 | 21,686 | 5,233 | 0.98 | 6,964 | 7,087 |
| V-38 | Vellore | 4.50 | 24,333 | 5,404 | 1.34 | 9,610 | 7,162 |
| V-39 | Vellore | 175.20 | 283,487 | 1,618 | 15.38 | 109,288 | 7,105 |
| V-40 | Vellore | 175.50 | 286,324 | 1,631 | 15.62 | 111,754 | 7,153 |
| V-41 | Vellore | 0.42 | 2,925 | 6,917 | 0.22 | 1,490 | 6,633 |
| V-42 | Vellore | 0.07 | 213 | 3,267 | 0.03 | 127 | 3,780 |
| V-43 | Vellore | 1.38 | 2,202 | 1,596 | 0.24 | 825 | 3,370 |
| V-44 | Vellore | 21.69 | 24,206 | 1,116 | 1.43 | 6,748 | 4,708 |
| V-45 | Vellore | 25.22 | 45,844 | 1,818 | 3.49 | 25,292 | 7,248 |
| V-46 | Vellore | 0.15 | 1,844 | 12,072 | 0.15 | 1,844 | 12,072 |
| V-47 | Vellore | 0.12 | 728 | 6,019 | 0.12 | 728 | 6,019 |
| V-48 | Vellore | 62.60 | 105,523 | 1,686 | 2.32 | 9,782 | 4,214 |
| V-49 | Vellore | 0.04 | 380 | 9,050 | 0.04 | 380 | 9,050 |
| V-50 | Vellore | 0.05 | 644 | 14,026 | 0.05 | 644 | 14,026 |
| V-51 | Vellore | 1.43 | 5,824 | 4,080 | 0.07 | 435 | 5,900 |
| V-52 | Vellore | 0.33 | 2,732 | 8,273 | 0.05 | 496 | 9,332 |
| V-53 | Vellore | 0.26 | 2,161 | 8,182 | 0.26 | 2,161 | 8,182 |
| V-54 | Vellore | 0.12 | 1,157 | 9,805 | 0.12 | 1,157 | 9,805 |
| V-55 | Vellore | 0.13 | 1,014 | 7,633 | 0.13 | 1,014 | 7,633 |
| V-56 | Vellore | 0.08 | 567 | 7,306 | 0.08 | 567 | 7,306 |
| V-57 | Vellore | 0.10 | 1,059 | 10,451 | 0.10 | 1,059 | 10,451 |
| V-58 | Vellore | 0.18 | 1,599 | 8,784 | 0.18 | 1,599 | 8,784 |
| V-59 | Vellore | 0.52 | 4,476 | 8,581 | 0.52 | 4,476 | 8,581 |
| V-60 | Vellore | 0.24 | 3,112 | 13,155 | 0.24 | 3,112 | 13,155 |
| V-61 | Vellore | 0.11 | 1,470 | 13,811 | 0.11 | 1,470 | 13,811 |
| V-62 | Vellore | 0.08 | 1,148 | 13,868 | 0.08 | 1,148 | 13,868 |
| V-63 | Vellore | 25.59 | 50,280 | 1,965 | 3.80 | 29,465 | 7,757 |
| V-64 | Vellore | 24.20 | 35,884 | 1,483 | 2.56 | 15,966 | 6,234 |
| V-65 | Vellore | 22.04 | 28,441 | 1,290 | 1.79 | 10,984 | 6,146 |
| V-66 | Vellore | 21.91 | 26,825 | 1,224 | 1.66 | 9,367 | 5,637 |
| V-67 | Vellore | 21.76 | 25,011 | 1,149 | 1.51 | 7,553 | 4,999 |
| V-68 | Vellore | 21.53 | 22,989 | 1,068 | 1.28 | 5,532 | 4,330 |
| V-69 | Vellore | 21.48 | 22,561 | 1,050 | 1.23 | 5,104 | 4,153 |
| V-70 | Vellore | 21.40 | 21,917 | 1,024 | 1.15 | 4,460 | 3,883 |
| V-71 | Vellore | 0.04 | 554 | 13,641 | 0.04 | 554 | 13,641 |
| V-72 | Vellore | 0.05 | 517 | 10,871 | 0.05 | 517 | 10,871 |
| V-73 | Vellore | 1.63 | 3,874 | 2,380 | 0.49 | 2,498 | 5,066 |
| V-74 | Vellore | 0.23 | 802 | 3,542 | 0.06 | 228 | 3,556 |
| V-75 | Vellore | 0.12 | 780 | 6,372 | 0.12 | 723 | 6,248 |
| V-76 | Vellore | 0.83 | 6,077 | 7,279 | 0.36 | 2,406 | 6,691 |
| V-77 | Vellore | 0.93 | 6,687 | 7,176 | 0.46 | 3,016 | 6,605 |
| V-78 | Vellore | 1.16 | 8,453 | 7,278 | 0.69 | 4,783 | 6,970 |
| V-79 | Vellore | 0.11 | 689 | 5,995 | 0.11 | 689 | 5,995 |
| V-80 | Vellore | 0.07 | 528 | 7,878 | 0.07 | 528 | 7,878 |
| V-81 | Vellore | 0.06 | 347 | 5,641 | 0.06 | 347 | 5,641 |
| V-82 | Vellore | 62.59 | 105,380 | 1,684 | 2.30 | 9,640 | 4,183 |
| V-83 | Vellore | 62.47 | 104,508 | 1,673 | 2.19 | 8,767 | 4,010 |
| V-84 | Vellore | 5.45 | 26,629 | 4,885 | 0.65 | 3,720 | 5,700 |
| V-85 | Vellore | 139.21 | 156,003 | 1,121 | 5.30 | 20,391 | 3,847 |
| V-86 | Vellore | 62.77 | 106,554 | 1,698 | 2.47 | 10,776 | 4,354 |
| V-87 | Vellore | 62.74 | 106,343 | 1,695 | 2.45 | 10,565 | 4,317 |
| V-88 | Vellore | 0.09 | 403 | 4,632 | 0.07 | 365 | 4,892 |
| V-89 | Vellore | 139.96 | 160,171 | 1,144 | 6.05 | 24,559 | 4,059 |
| V-90 | Vellore | 139.75 | 158,690 | 1,136 | 5.84 | 23,078 | 3,954 |
| V-91 | Vellore | 75.16 | 40,103 | 534 | 2.02 | 3,941 | 1,954 |
| V-92 | Vellore | 75.03 | 39,504 | 527 | 1.89 | 3,379 | 1,783 |
| V-93 | Vellore | 0.21 | 1,442 | 6,977 | 0.02 | 63 | 3,595 |
| V-94 | Vellore | 0.05 | 298 | 6,393 | 0.05 | 298 | 6,393 |
| V-95 | Vellore | 4.57 | 25,148 | 5,501 | 1.41 | 10,425 | 7,393 |
| V-96 | Vellore | 147.03 | 210,891 | 1,434 | 9.95 | 60,557 | 6,084 |
| V-97 | Vellore | 147.08 | 211,438 | 1,438 | 10.00 | 61,103 | 6,109 |
| V-98 | Vellore | 174.54 | 277,750 | 1,591 | 14.96 | 104,723 | 6,998 |
